# Effects of racial/ethnic disparities in healthcare utilization on antibiotic use, United States, 2016/2018

**DOI:** 10.1101/2021.12.09.21266965

**Authors:** Scott W. Olesen, Sanjat Kanjilal, Stephen M. Kissler, Daphne S. Sun, Yonatan H. Grad

**Affiliations:** Department of Immunology and Infectious Diseases, Harvard Chan School of Public Health, Boston, MA, USA; Department of Population Medicine, Harvard Medical School & Harvard Pilgrim Healthcare Institute, Boston, MA, USA

**Keywords:** antibiotics, race/ethnicity, equity, healthcare utilization

## Abstract

White Americans make more office-based and emergency department visits per capita than other races/ethnicities, but the proportion of visits during which antibiotics are administered or prescribed is similar across races/ethnicities. Racial/ethnic disparities in antibiotic use may be due more to disparities in healthcare access and utilization than to prescriber behavior.

## INTRODUCTION

Antibiotics are an important therapy for treating infectious disease, but use of antibiotics carries risk of negative effects, like adverse events or antibiotic resistance. Avoiding antibiotic overuse through antibiotic stewardship helps avoid unnecessary and inappropriate antibiotic use. However, underuse of antibiotics, when a person has an infection for which antibiotics are medically recommended but the person does not receive the antibiotic, is also problematic [1]. Even for conditions like sore throat or otitis media, which typically resolve without complications, failure to use antibiotics can sometimes lead to substantial morbidity [2].

Disparities in antibiotic use rates, when one population uses more or less antibiotics than another group, are a likely indicator of antibiotic overuse or underuse [1,3]. Many studies have shown that prescribing practice varies by patient race/ethnicity, usually with whites more likely to receive antibiotics or broader-spectrum antibiotics relative to people of other races [1,4–15], although other studies have found no effect [16–23] or even the opposite effect [24,25]. The most common topic of these studies is antibiotic prescribing for acute upper respiratory infections in children. A typical finding is that if two children, one white and one Black, present with identical symptoms, the white child is more likely to receive antibiotics.

Studies of antibiotic prescribing per healthcare visit, while important, do not address all the factors that could lead to disparities in antibiotic use. First, rates of bacterial infections may vary between populations. Second, access to healthcare and healthcare seeking behavior may differ across populations. In other words, the probability that someone with an infection presents at a healthcare visit may vary by population. Third, different populations may be more or less likely to receive antibiotics when they present at a healthcare visit. Studies of prescribing per visit only address the third step, but previous work suggests that geographical differences in antibiotic use rates are due more to differences in healthcare access and utilization than to prescribing practice [26] and that temporal trends in racial disparities in antibiotic use among children are due more to changes in healthcare utilization than to changes in prescribing practice [7].

Here, we use a nationally representative survey of healthcare visits to assess the relative importance of healthcare utilization and prescribing practice on antibiotic use by race/ethnicity. We hypothesized that differences in healthcare utilization, measured as per capita rates of ambulatory care visits, and differences in prescribing practice, measured as the probability that an antibiotic will be used given that certain diagnoses are present, both contribute to any racial/ethnic differences in antibiotic use.

## METHODS

We analyzed data from the National Ambulatory Medical Care Survey (NAMCS) and National Hospital Ambulatory Medical Care Survey (NHAMCS), two nationally representative samples used to characterize antibiotic prescribing practice [27,28]. Each survey is a sample of office-based and emergency department visits with associated patient demographics, diagnosis codes, and prescriptions as provided by physicians or hospital staff. Missing race/ethnicity data are imputed using a model developed by NAMCS/NHAMCS staff [28]. We used the two most recent years with data from both surveys, 2016 and 2018. NHAMCS from 2017 data were not available [29].

Each healthcare visit was classified as antibiotic-appropriate, potentially antibiotic-appropriate, or antibiotic-inappropriate by a two-step process [30]. First, using previously established categorizations of the ICD-10 diagnosis codes, each diagnosis code associated with each visit was classified as always antibiotic-appropriate, sometimes antibiotic-appropriate, or never antibiotic-appropriate. Second, each visit was classified based on its diagnoses. A visit with at least one always-appropriate diagnosis was classified as antibiotic-appropriate. Remaining unclassified visits with at least one sometimes-appropriate code were classified as potentially antibiotic-appropriate. Remaining unclassified visits with at least one never-appropriate code were classified as antibiotic-inappropriate.

Separately, each visit was classified as a visit with antibiotics if any of the prescribed or administered medications included oral antibiotics [27].

Note that all visits are classified according to their associated diagnoses, regardless of whether antibiotics were prescribed or not. Thus, most visits are considered antibiotic-inappropriate, not because patients are seeking antibiotics for antibiotic-inappropriate conditions, but rather because most healthcare visits are not made for the purpose of treating bacterial infections.

Differences in rates were assessed using *t*-tests, with *p* < 0.01 considered statistically significant, as recommended in the NHAMCS documentation [28]. Differences in proportions were assessed using *χ*^2^ tests. All analyses were performed using R (version 4.0.5) [31], with the *survey* package (version 4.0) [32] used to account for the complex survey design in variance estimation. The population denominators for each race/ethnicity provided in the NAMCS/NHAMCS documentation, which are drawn from US Census estimates, were used for population-based rate estimates. Code to reproduce these analyses is available at GitHub (doi: 10.5281/zenodo.6233588).

## RESULTS

During 2016 and 2018, rates of physician office and hospital emergency room visits varied by race/ethnicity. Non-Hispanic whites made the highest rate of visits, followed by non-Hispanic Blacks, Hispanics, and non-Hispanic people of multiple or other races (Figure 1a, Table S1). The difference in rates of visits was mostly due to different rates of visits with no associated diagnoses for which antibiotics would be appropriate or sometimes appropriate (*t*-tests, Figure 1a, Table S1).

**Figure 1.**
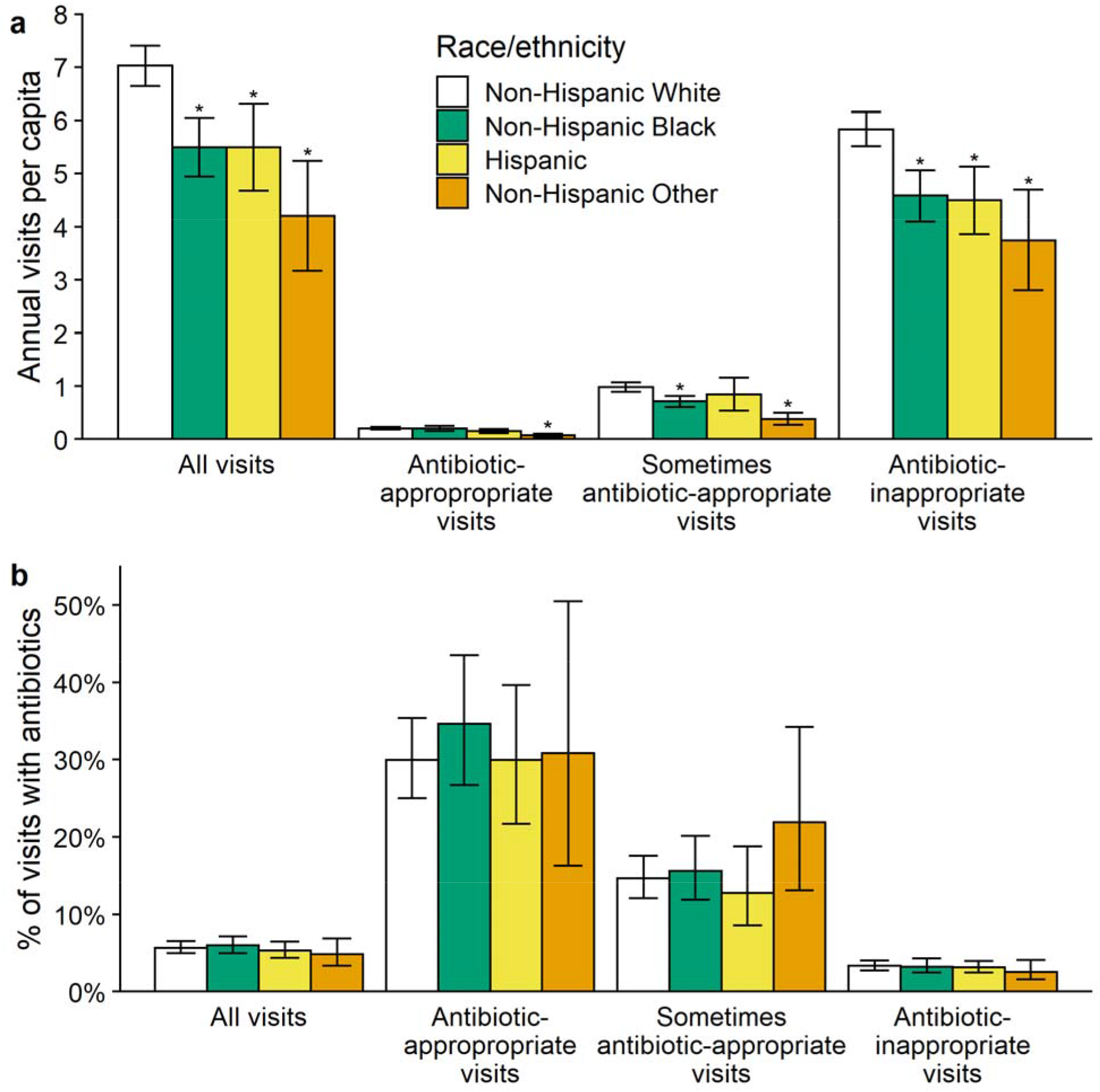
(a) Annual office-based and emergency department visits per capita, stratified by patient race/ethnicity and by antibiotic-appropriateness of the diagnoses recorded for the visit. Error bars show 95% confidence intervals. *: *t*-test, *p* < 0.01. (b) Proportion of visits with antibiotics administered or prescribed.

Non-Hispanics whites had a higher rate of visits with antibiotics administered or prescribed compared to non-Hispanic people of multiple or other races, but differences in rates of visits with antibiotics were not statistically significant for non-Hispanic whites compared to non-Hispanic Blacks or to Hispanics (Table S2).

The proportion of visits during which antibiotics were prescribed or administered (5.5%, 95% CI 4.9% to 6.0%) did not vary statistically significantly by race/ethnicity (*p* = 0.57, *χ*^2^ test), even when stratifying by the antibiotic-appropriateness of the diagnoses associated with the visit (*p* > 0.05, *χ* ^2^ tests; Figure 1b, Table S3).

## DISCUSSION

In this analysis of nationally representative survey data, whites made more healthcare visits per capita. However, there was no statistically significant difference across races/ethnicities in the proportion of visits that included antibiotics, even when stratifying by whether antibiotics were appropriate for the visit. These results suggest that differences in healthcare utilization are at least as important as per-visit prescribing practice to disparities in antibiotic use. For example, whites had a 27% higher rate of healthcare visits than Blacks (7.0 vs. 5.5 annual visits per capita), but the point estimate in the proportion of visits with antibiotics was 6% lower for whites (5.7% vs. 6.0% of visits with antibiotics prescribed). Thus, any difference in the overall rate of antibiotic use between whites and Blacks is likely more attributable to differences in the number of opportunities for antibiotics to be prescribed rather than to the probability that an antibiotic will be prescribed at a visit.

This study’s primary strength is its use of a nationally representative survey. Its most important limitation is that the sampling frame is healthcare visits. A survey of visits can estimate visits by race/ethnicity and the proportions of visits with antibiotics, but it cannot address the proportion of infections that are treated with antibiotics because not every infection that merits antibiotics results in a visit. Thus, without knowing the underlying rates of disease, we cannot determine whether patients of different races/ethnicities with antibiotic-treatable infections are more or less likely to present for treatment [1]. For example, if non-whites had higher underlying rates of bacterial infections, then whites’ approximately equal rates of antibiotic-appropriate visits actually represent a disparity in healthcare access or healthcare seeking.

This study has other important limitations. First, these national surveys did not have sufficient statistical power to characterize any differences across races/ethnicities in the probability of per-visit antibiotic use. For example, the point estimate for the proportion of Blacks’ antibiotic-appropriate visits that had antibiotics was higher than that for whites, but the confidence intervals were wide and overlapping (Figure 1b, Table S3). Second, these surveys do not include every kind of visit type and exclude urgent care, telephone or telehealth contact, hospital outpatient departments, Veterans Administration hospitals, and inpatient care. People of different races/ethnicities seek care in different healthcare contexts [7] and so these results are necessarily incomplete. Third, there may be systematic biases in the diagnosis codes that prescribers assign to people of different races with the same underlying disease [1]. Finally, the available data cannot assess the effect of the Covid-19 pandemic on antibiotic prescribing patterns.

Despite these limitations, these results point to two important components for addressing equity in the quality of care for bacterial infections: first, characterizing racial/ethnic differences in underlying rates of antibiotic-treatable infections, and second, mitigating barriers to healthcare access and utilization.

## Data Availability

Source data are openly available from the National Center for Health Statistics (https://ftp.cdc.gov/pub/Health_Statistics/NCHS/Datasets, https://www.cdc.gov/nchs).

https://www.cdc.gov/nchs

## STATEMENTS

### Patient content

This work is not human subjects research.

